# Exploiting Zeta potential of air borne pathogenic bacteria for effective air decontamination

**DOI:** 10.1101/2025.02.08.25321938

**Authors:** Arun Sai Kumar Peketi, SVL Sai Kushal Kumar, P Kennedy Kumar, Bulagonda Eswarappa Pradeep

**Author notes:** Both the authors contributed equally (Arun Sai Kumar Peketi, SVL Sai Kushal Kumar).

## Abstract

**Background:** Airborne microorganisms contribute significantly to hospital-associated infections (HAIs), particularly causing respiratory tract infections (RTIs). Multi-drug resistant (MDR) airborne pathogens pose a global clinical threat, challenging existing air decontamination technologies. While current methods effectively trap microbes, they often lack microbicidal properties. Exploitation of Zeta potential of microbes which are naturally charged could be a potential strategy for air decontamination, independent of their antimicrobial resistance. In this study, we tested the efficacy of ZeBox technology which relies on zeta potential to trap and eliminate airborne pathogens such as clinically isolated, MDR *Klebsiella pneumoniae, Acinetobacter baumannii, Pseudomonas aeruginosa, Escherichia coli* of respiratory origin and a non RTI isolate *Staphylococcus aureus*.

**Methods:** Five MDR bacterial pathogens which include *K. pneumoniae* (2 nos), *A. baumannii, P. aeruginosa*, and *E. coli* from patients with RTI and one pus isolate of *S. aureus* were included in the study. These isolates were aerosolized in a certified BSL-2 setting containing a ZeBox-powered Air Sterilization device. Viable bacteria were enumerated before and after exposure to the Zebox powered device. Clinical and laboratory isolated bacterial strains were assessed for zeta potential variations to understand its association with antibiotic resistance and pathogenicity.

**Results:** Our analyses revealed that Zeta potential is species specific, but independent of the pathogenicity and antibiotic susceptibility of the tested bacteria. Exposure to Zebox powered device for 5 mins resulted in a remarkable decline of microbial load with a minimum 5 log reduction (99.999%) of all the tested isolates, irrespective of their species. Zeta potential measurements further indicated a consistent killing mechanism of MDR pathogens by Zebox technology.

**Conclusions:** The current study underscores the reliability of zeta potential based air decontamination strategies for potential elimination of diverse, air borne MDR bacteria in healthcare settings.

**Highlights:**

- Zeta potential value of bacteria is not influenced by AST or pathogenicity of bacteria.
- Zeta potential driven device traps and kills airborne MDR bacteria with 99.999% efficiency.
- The efficacy of ZeBox technology is consistent across pathogens.

## 1. Introduction

Respiratory Tract Infections (RTIs) are among the top four causes of morbidity and mortality worldwide, accounting for 6% of global disease burden [1] and 3%-5% of global adult mortality [2]. In 2019 alone, lower RTIs were responsible for more than 1.5 million deaths [3]. The leading pathogens behind these fatalities include antibiotic-resistant strains of *Escherichia coli, Staphylococcus aureus, Klebsiella pneumoniae, Streptococcus pneumoniae, Acinetobacter baumannii*, and *Pseudomonas aeruginosa*. These pathogens were reported to account for approximately one million deaths directly attributable to antimicrobial resistance (AMR) and an additional 3.5 million deaths associated with AMR [3].

India alone accounts for 23% of the global pneumonia burden with high mortality rates [4]. Community-acquired lower respiratory tract infections (LRTIs) were reported to be a common cause of death among older adults [5]. The spectrum of bacterial pathogens which includes *Klebsiella pneumoniae, Staphylococcus aureus* and *Escherichia coli*, along with their frequencies were reported as the primary causative agents of RTIs [6].

The emergence of Multi-Drug Resistance (MDR) among bacteria causing RTI undermines the efficacy of available treatment options and leads to increased mortality rates due to treatment failures [3].

All microorganisms, pathogenic or non-pathogenic, possess negatively charged outer membranes [7]. Zeta potential is a representative of the surface potential of the bacteria which arises due to the distribution of charges on their cell surfaces. These surface charges primarily arise from the molecules and ions on the bacterial cell wall and membrane. In Gram-positive bacteria, the thick peptidoglycan layer contains teichoic and lipoteichoic acids, contributing negatively charged groups (such as phosphate groups) to the cell surface [7]. In Gram-negative bacteria, an outer membrane containing lipopolysaccharides (LPS) contributes to the overall negative charge due to phosphate and carboxyl groups.

Additionally, bacteria may have various surface proteins, polysaccharides, and glycoproteins contributing to their surface charge. This electrostatic potential plays a significant role in how bacteria interact with their surroundings, including their adhesion to surfaces and aggregation behaviour.

Previous reports indicated that ZeBox air sterilization technology exploits the bacterial zeta potential to trap and kill microorganisms from ambient air [7, 8]. It employs a non-ionizing electric field that manipulates the movement of these charged entities (microorganisms) and traps them onto a three-dimensional, specially engineered surface coated with Quaternary Ammonium Salt (QAS). The killing of the trapped microorganisms is mediated by the synergistic effect of the electric field and QAS [7].

In this study, the zeta potential of multiple MDR bacteria both from respiratory and non-respiratory origin were measured and were above −29mV. We hypothesised that Zebox technology which exploits zeta potential can effectively eliminate air borne bacteria irrespective of their classification and antibiotic resistance. We challenged the ZeBox technology powered device with aerosolized MDR isolates collected from clinical settings.

## 2. Methods

### 2.1 Sample collection, isolation and Antibiotic Susceptibility Testing

Respiratory Tract Infection (RTI) and Pus isolates collected from Sri Ramachandra Institute of Higher Education and Research, Chennai, India were shipped as agar stabs to the Antimicrobial Resistance Laboratory (AMR-LAB) at SSSIHL, India. The identity of each organism was confirmed by Vitek-2. Antibiotic Susceptibility Testing (AST) was performed using the Kirby-Bauer Disk Diffusion (KBDD) method against various commonly used antimicrobial agents as per CLSI guidelines [9]. Five RTI and one pus isolate confirmed to be multi-drug resistant (MDR) were chosen for the study. *Escherichia coli* ATCC 35218, *Pseudomonas aeruginosa* ATCC 27853, *Staphylococcus aureus* ATCC 25923 and *Klebsiella pneumoniae* ATCC 700603 were used as controls for AST and zeta potential measurements.

### 2.2 Growth curve analysis and Measurement of zeta potential

Optical Density (OD) based growth curves were plotted for all the selected isolates. Briefly, overnight culture of 1ml was used to inoculate each of the three flasks of 100ml fresh (Miller Luria Bertani Broth (LB Broth, Himedia, Mumbai, M1245). OD at 600nm was recorded every hour using the UV-visible spectrophotometer (Eppendorf Biospectrophotometer Kinetic, Eppendorf AG, Germany). After determining the growth kinetics of all isolates, 1ml aliquot was taken from the culture at the mid-log phase for every isolate. The 1ml aliquot was serially diluted in sterile 0.05X PBS pH 7.2 to attain a final concentration of ~10^6 cfu/ml. The final aliquot was centrifuged at 25°C at 3,000 G. The supernatant was removed, and the pellet was washed with sterile 0.05X PBS, pH 7.2, multiple times. The final washed pellet was resuspended using sterile 0.05X PBS, and the zeta potential was measured using the Anton Paar Litesizer 500 instrument, with sterile 0.05X PBS pH 7.2 as a blank.

### 2.3 Sample preparation for aerosolization

A single pure colony of the bacteria was used to inoculate a 3ml Miller Luria Bertani Broth (LB Broth, Himedia, Mumbai, M1245) and was incubated for 12 hours at 37°C at 180 rpm. 1 ml of this growth culture was used as a seed to inoculate 100 ml of LB broth and was incubated for 12 hours at 37^°^C at 180 rpm. After incubation, the culture was placed in 50ml sterile tubes and centrifuged at 25^°^C at 3,000g. The supernatant was discarded and the pellet was washed with 5 ml of sterile 0.05 X PBS pH 7.2. The final washed pellet was resuspended in 5ml of sterile 0.05 X PBS pH 7.2 and was loaded into the nebulizer for aerosolization.

### 2.4 Test chamber setup for aerosolization of bacteria

The test chamber was set up inside a Biosafety level 2 laminar flow hood coupled with an incinerator and exhaust, which was in operation during the entire experiment. An air-sealed test chamber of dimensions 3.5 ft. x 2.5 ft. x 2ft. was built with sampling and nebulization ports. A six-jet Collison nebulizer (BGI Inc.) was used in this study, which produces aerosols of a mean diameter of 2-5um. Dry air from a compressed air cylinder at a pressure of 10 psi was used to operate the nebulizer. The nebulized bacteria were collected at different time intervals onto an MCE membrane of 0.22µm pore size (Cobetter, MFMCE-2225), and assembled within a filter unit (Tarsons 521090) connected to a vacuum pump. The collection was performed for 10 min at 0.02MPa.

### 2.5 Device description

A miniature version of the ZeBox technology was used during this study, where 5 electrode plates were connected parallel to the airflow. A 30CFM fan was used to pull the contaminated air from the chamber and pass it through the channels formed by the electrode plates. The electrode plates are covered with a 3-dimensional surface coated with QAS. The plates are connected to a non-ionizing electric field, generating a potential difference of 2.4-4.2kVDC/cm. The electrode plates had a dimension of 15cm x 6cm with a thickness of 1.6mm. (Supplementary Figure 1)

**Supplementary Figure 1:**
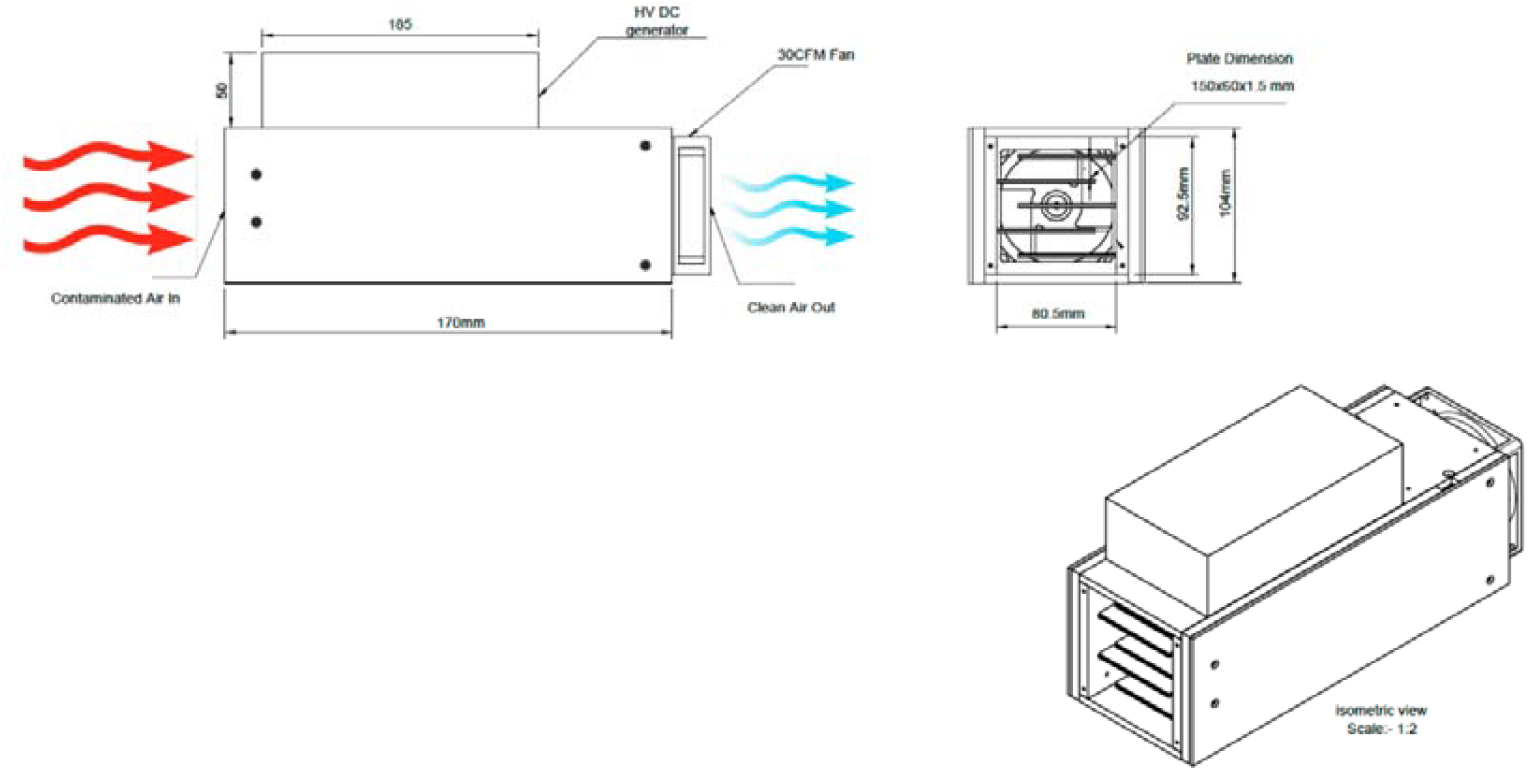
Design blueprint of ZeBox technology in a miniature version.

### 2.6 Enumeration of the collected microorganisms

The MCE membrane was used to collect the aerosolized microorganisms from the test chamber and was suspended in 5 ml of sterile 0.05 X PBS pH 7.2. The suspension was then serially diluted using sterile 0.05 X PBS pH 7.2, and appropriate dilutions were plated on Luria Bertani Agar (LB agar, Hi-Media, Mumbai, M1151). The agar plates were incubated for 24 hours, and the individual colonies were enumerated.

## 3. Results

### 3.1 Selection of isolates

A total of 17 isolates were identified and tested for their antibiotic susceptibility, along with standard lab strains. Six clinical isolates were selected based on their Antibiotic Susceptibility Testing (AST) patterns. These included two isolates of *Klebsiella pneumoniae* one isolate each of *Pseudomonas aeruginosa, Staphylococcus aureus, Escherichia coli* and *Acinetobacter baumannii*. Five of these six organisms were isolated from the sputum of RTI patients. Methicillin-resistant *Staphylococcus aureus* was a pus isolate from a non-RTI patient. *Escherichia coli* ATCC 35218, *Pseudomonas aeruginosa* ATCC 27853, *Klebsiella quasipneumoniae* ATCC 700603 and *Staphylococcus aureus* ATCC 25923 were used as controls for AST testing and zeta potential measurements (Table 1).

**Table 1:** Identity and AST of six isolates selected for assessing the efficacy of ZeBox, along with the AST of available laboratory strains and zeta potential values.

| Isolate |  | GE5133 | GE5114 | GR4696 | GR4698 | GR4761 | PS36 | EC | PA | KP | SA |
| --- | --- | --- | --- | --- | --- | --- | --- | --- | --- | --- | --- |
| Antibiotic Class | Antibiotic | *Escherichia coli* | *Acinetobacter baumannii* | *Pseudomonas aeruginosa* | *Klebsiella pneumoniae* | *Klebsiella pneumoniae* | *Staphylococcus aureus* | *Escherichia coli* ATCC 35218 | *Pseudomonas aeruginosa* ATCC27853 | *Klebsiella quasipneumoniae* ATCC700603 | *Staphylococcus aureus* ATCC25923 |
| Penicillins | Ampicillin | R | - | - | R | R | - | S | S | R | S |
|  | Penicillin | - | - | - | - | - | R | S | S | R | S |
|  | Piperacillin tazobactam | - | R | - | - | - | - | S | S | R | S |
|  | Oxacillin | - | - | - | - | - | R | S | S | R | S |
| Cephalosporins | Cefepime | R | R | R | R | S | - | S | S | S | S |
|  | Cefazolin | R | - | - | R | R | - | S | S | R | S |
|  | Cefuroxime | R | - | - | R | R | - | S | S | R | S |
|  | Ceftazidime | R | R | R | R | S | - | S | S | R | S |
|  | Cefoxitin | S | - | - | R | S | - | S | S | R | S |
|  | Cefotaxime | R | R | - | R | R | - | S | S | R | S |
|  | Cefoperzone-sulbactam | S | R | - | R | R | - | S | S | R | S |
|  | Cefixime | R | R | R | R | S | - | S | S | R | S |
| Carbapenems | Imipenem | S | R | R | R | S | - | S | S | S | S |
|  | Ertapenem | - | - | - | - | - | - | S | S | S | S |
|  | Meropenem | S | R | R | R | S | - | S | S | S | S |
| Aminoglycoside | Amikacin | S | R | R | S | S | - | S | S | S | S |
|  | Gentamicin | S | R | S | S | S | R | S | S | S | S |
|  | Tobramycin | - | - | R | - | - | - | S | S | S | S |
| Fluoroquinolone | Levofloxacin | - | - | - | - | - | R | S | S | S | S |
|  | Norfloxacin | R | - | R | R | S | - | S | S | S | S |
|  | Ciprofloxacin | R | - | R | R | S | R | S | S | S | S |
| Tetracycline | Tigecycline | S | S | - | S | S | S | S | S | S | S |
|  | Minocycline | S | S | R | S | S | - | S | S | S | S |
|  | Tetracycline | R | - | - | S | S | S | S | S | S | S |
| Nitrofurans | Nitrofurantoin | S | - | - | R | S.5 | S.5 | S | S | S | S |
| DHFR Inhibitor | Co-trimoxazole | R | R | - | R | S | R | S | S | S | S |
| Category |  | MDR | MDR | MDR | XDR | MDR | MDR | AS | AS | AS | AS |
| Zeta Potential (mV) |  | -33.4 | -29.8 | -35.6 | -37.1 | -37.2 | -36.4 | -44.2 | -43.8 | -38.2 | -33.5 |

### 3.2 Measuring Zeta potential of selected MDR and reference strains

The zeta potential values of the selected isolates ranged between −29mV and −44.2mV. among clinical isolates, *Acinetobacter baumannii* (GE5114) had the lowest zeta potential (−29.8mV), and *Klebsiella pneumoniae* (GR4761) had the highest zeta potential of −37.2mV. The observed zeta potentials for the control strains of *Escherichia coli, Pseudomonas aeruginosa, Klebsiella quasipneumoniae* and *Staphylococcus aureus* were −44.2mV, −43.8mV, −38.2mV and −33.5mV respectively. The zeta potential values of the tested isolates are mentioned in Table 1, and the zeta potential distribution graphs are presented in Figure 1.

**Figure 1:**
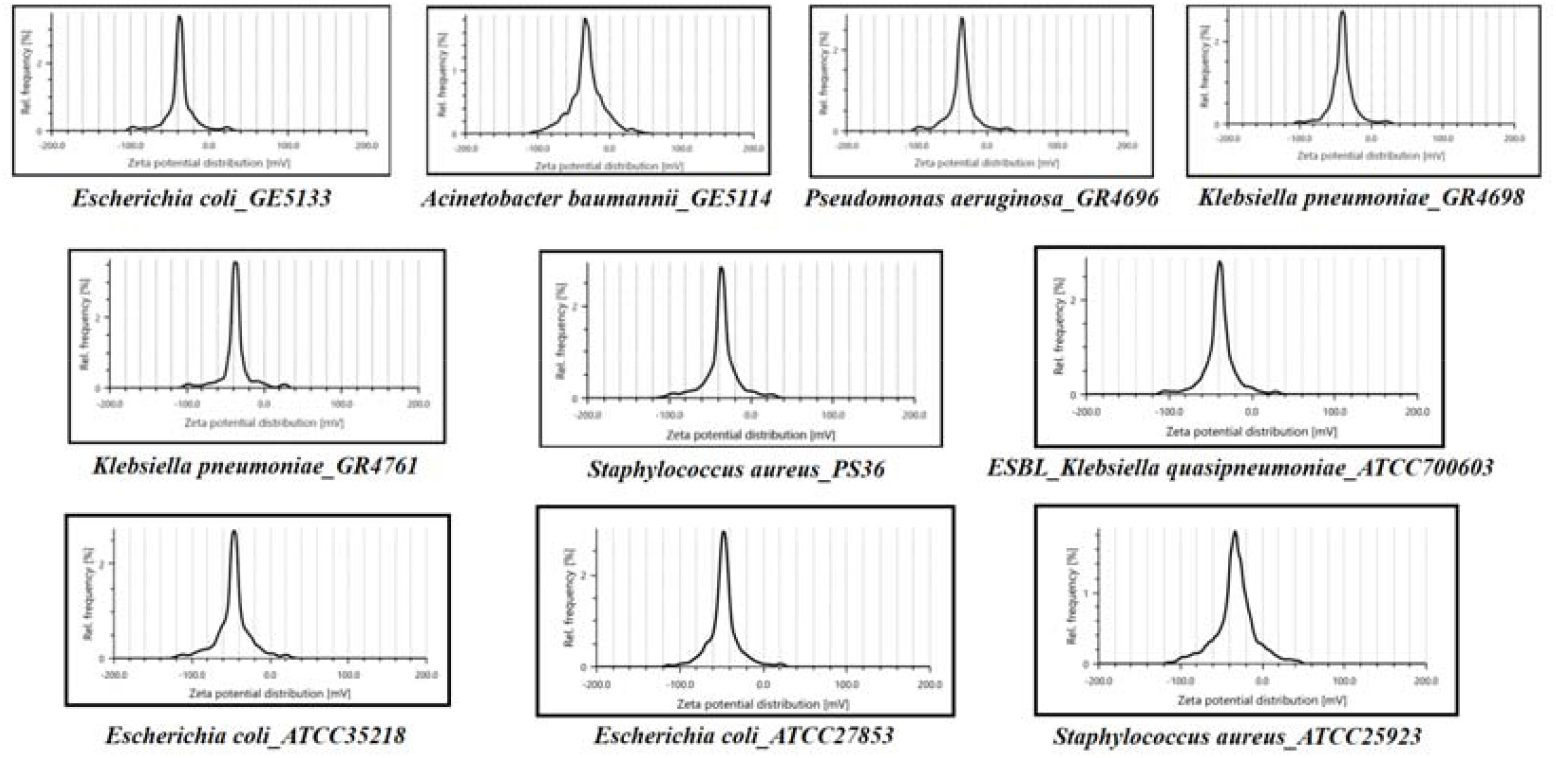
Zeta potential for the tested isolates and available reference strains. The zeta potential values observed for the clinical isolates were relatively lower than their corresponding laboratory strains.

### 3.3 ZeBox technology eliminates airborne MDR strains

The mean baseline airborne bacterial load generated by nebulization was initially determined to be around 10^9^ CFU. Upon operation of Zebox technology, the mean airborne bacterial load inside the test chamber after one minute was found to be around 10^7^ CFU (reduction of 98% or 1.8log_10_), around 10^6^ CFU (reduction of 99.9% or 3log_10_) after three minutes of operation and around 10^3^ CFU (reduction of 99.9999% or 6log_10_) after 5 minutes of operation. The airborne microbial elimination rates for individual multidrug-resistant isolates are presented in Figure 2 and Table 2, demonstrating the efficacy of ZeBox in rapidly and effectively eliminating airborne pathogens.

**Table 2:**
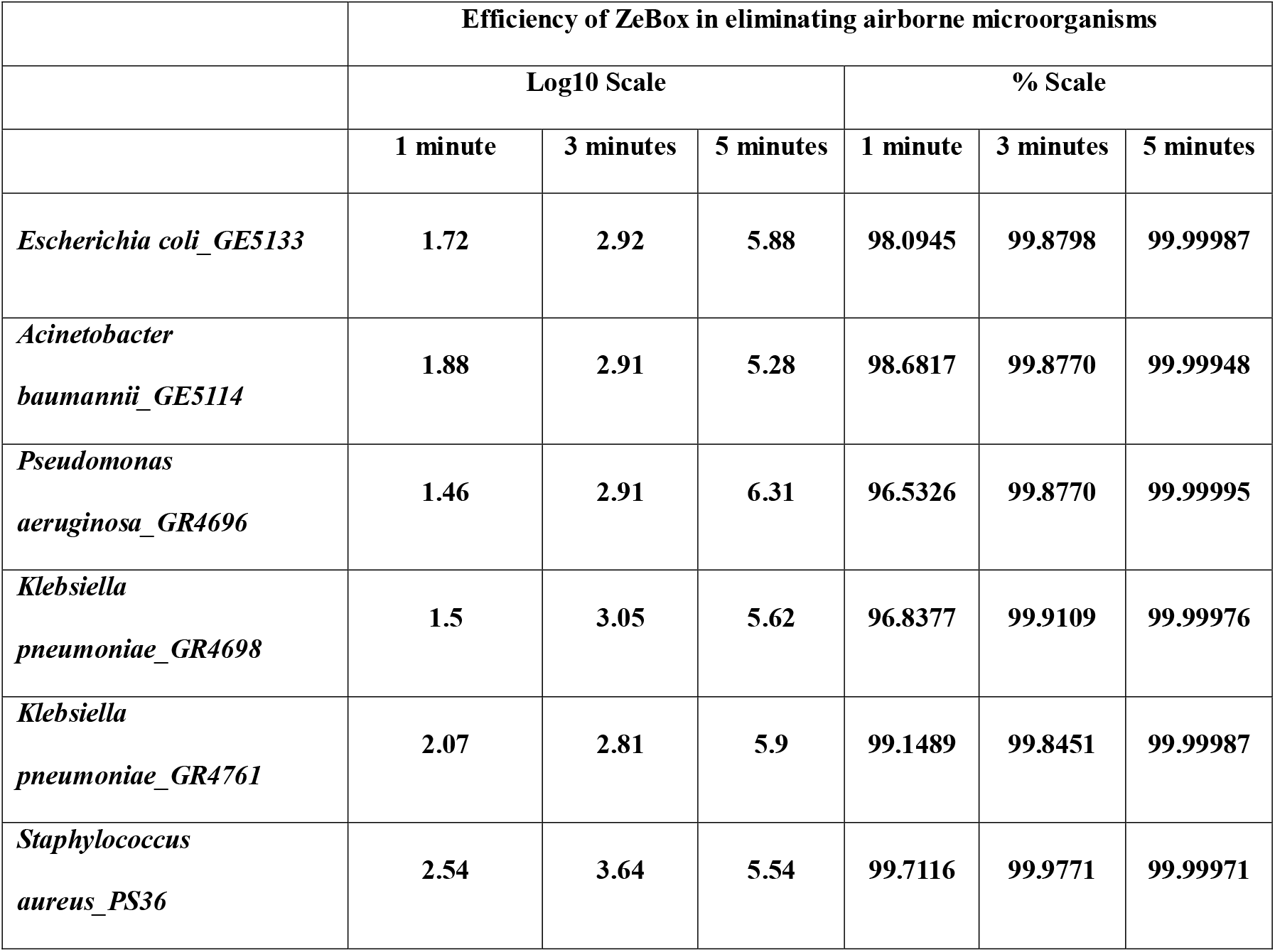
Comparative analysis of the airborne microbial elimination rate using two standard measuring scales.

**Figure 2:**
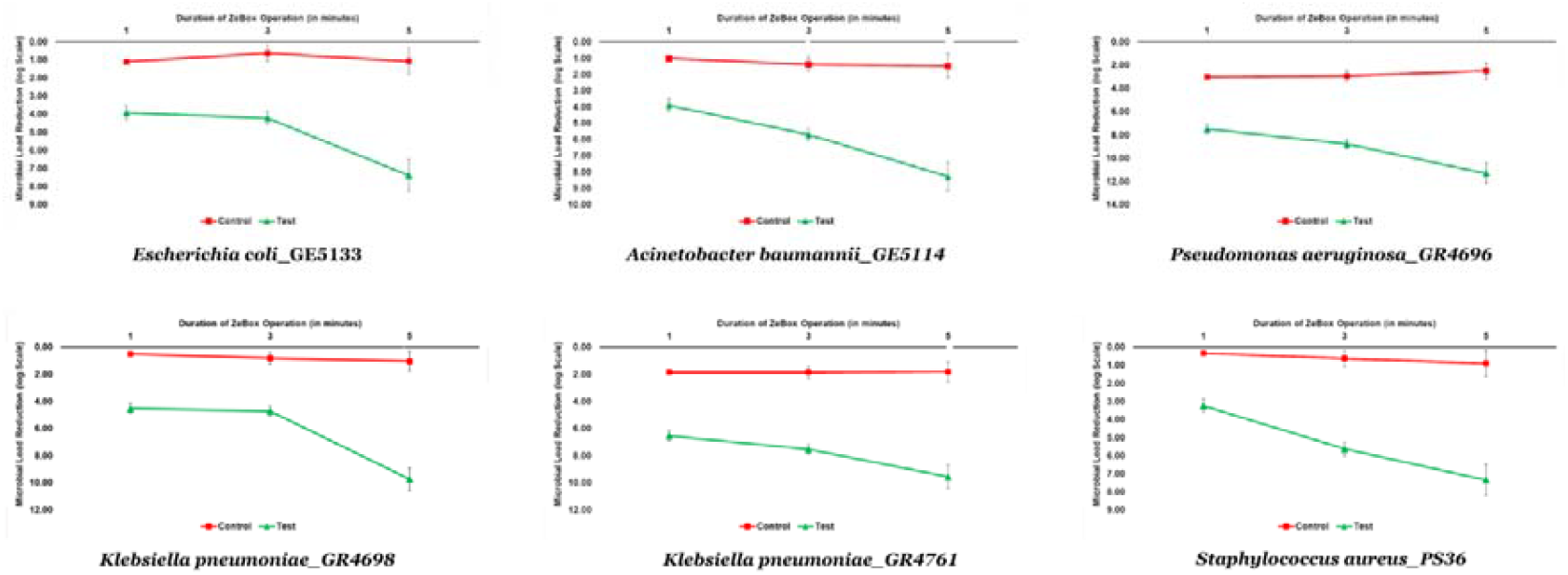
Airborne microorganisms elimination rate analysis for the six tested MDR isolates, when ZeBox device was not operated (red), vs. operation of ZeBox device for a definite period of time, (One minute, three minutes and five minutes). As observed, the microbial load inside the chamber settled down by ~1 log_10_ when the chamber was undisturbed; when ZeBox was operated, the microbial reduction was ~ ~6 log_10,_ indicating a 99.9999% reduction in airborne MDR strains under a controlled environment.

## 4. Discussion and conclusion

The present study underlines the utility of zeta potential in eliminating MDR bacterial pathogens. All the tested pathogens were part of the WHO list of priority pathogens. There is an unmet need for infection control related to airborne MDR pathogens. The current study indicates that zeta potential based Zebox technology can efficiently aid in curbing the spread of airborne pathogens thereby minimizing an individuals’ exposure to harmful microbes. Measurement of zeta potential for the test isolates was found to be ideal when 10^6^ cfu/ml of bacteria were tested. At this concentration, a single peak of zeta potential was observed, indicating the uniformity of the suspended particles/cells. Our analyses indicate consistency in ZeBox kill kinetics which relies solely on the zeta potential of the isolate. It was observed that the zeta potential of the isolate is independent of the bacterial classification and antibiotic resistance. Hence, zeta potential could be leveraged as a key factor in developing therapeutic strategies that can be effective against a plethora of MDR bacteria.

## Data Availability

All data produced in the present work are contained in the manuscript

## Acknowledgements

The authors acknowledge the support of Dr. Santanu Datta and Dr. Arindam Ghatak, Biomoneta Private Limited, Bangalore for providing access to Zebox powered instrument; UGC-SAP-DRS-III, DST-FIST and DBT-BIF, Govt. of India for the infrastructural support to the Department of Biosciences, SSSIHL, Prasanthi Nilayam and ICMR-SRF from Govt. of India to ASKP. BEP received extramural Research support from ICMR, Govt. of India. (Project ID: 2020e5867; OMI/27/2020-ECD-I and AMR/Adhoc/281/2022-ECD-II).

## Author Contributions

BEP designed the study, KPK provided the clinical isolates, ASKP and SVLSKK conducted the experiments and prepared the draft manuscript. BEP reviewed and prepared the final version of the manuscript.

## Ethical statement

This study was conducted following approval from the Institutional Ethics Committee (IEC) of Sri Sathya Sai Institute of Higher Learning (SSSIHL/IEC/PSN/BS/2014/03) and Sri Ramachandra Institute of Higher Education and Research, Porur, Chennai (CSP-III/24/SEP/11/393) in accordance with the ethical standards of the Declaration of Helsinki.

