## Supplementary figures and images for "Exploiting Zeta potential of air borne pathogenic bacteria for effective air decontamination"

### Supplementary Figure 1

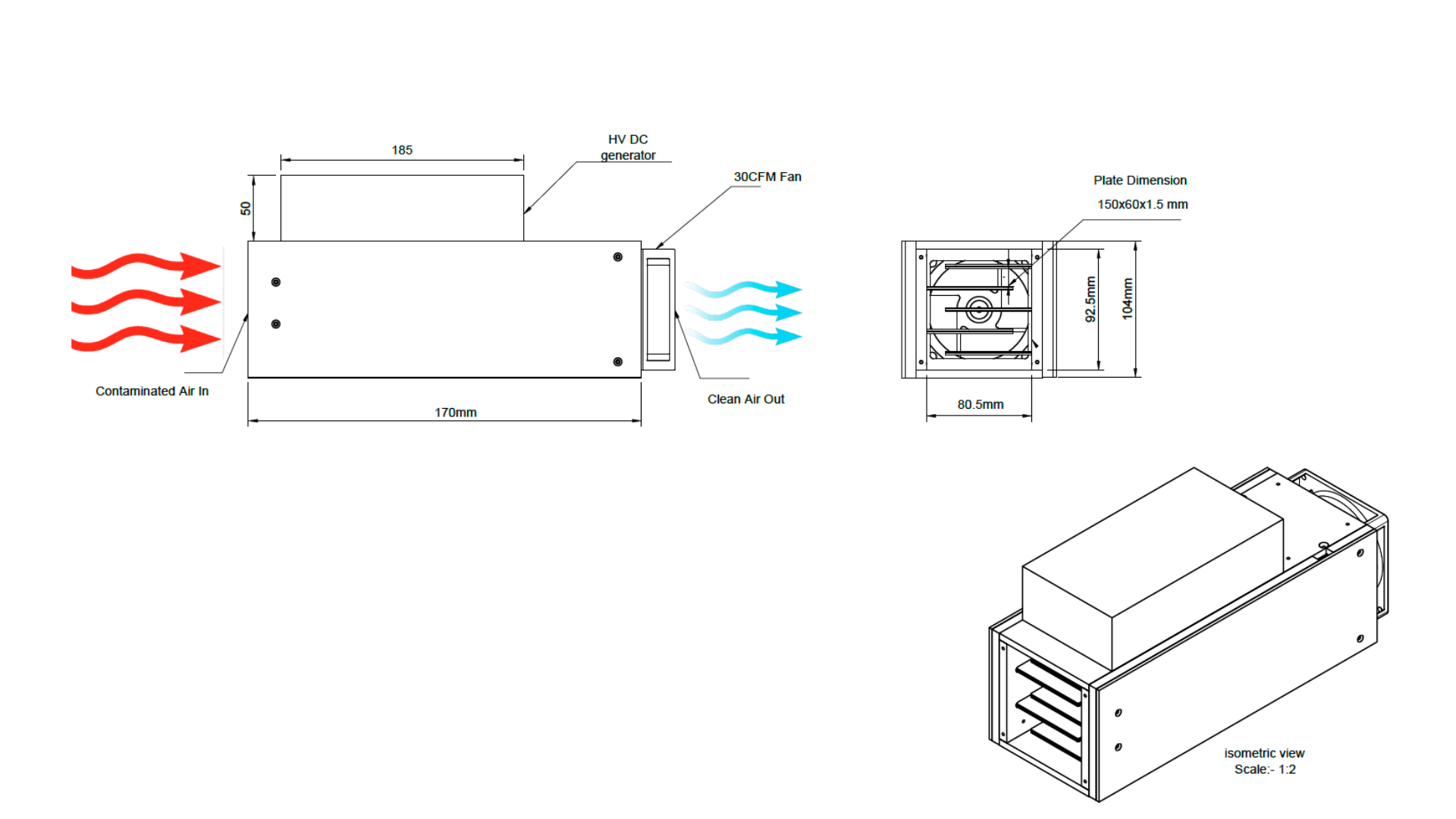
